# Estimation of the infection attack rate of mumps in an outbreak among college students using paired serology

**DOI:** 10.1101/2023.09.12.23295419

**Authors:** Michiel van Boven, Jantien A. Backer, Irene Veldhuijzen, Justin Gomme, Rob van Binnendijk, Patricia Kaaijk

**Affiliations:** Centre for Infectious Disease Control, National Institute for Public Health and the Environment, Bilthoven, the Netherlands; Julius Center for Health Sciences and Primary Care, University Medical Center Utrecht, Utrecht University, Utrecht, the Netherlands; Department of Epidemiology and Social Medicine, University of Antwerp, Antwerp, Belgium; NHS Scotland, Edinburgh, Scotland

## Abstract

Mumps virus is a highly transmissible pathogen that is effectively controlled in countries with high vaccination coverage. Nevertheless, outbreaks have occurred worldwide over the past decades in vaccinated populations. Here we analyse serological data from outbreaks of mumps virus genotype G among college students in the Netherlands over the period 2009-2012. To identify infections in the presence of preexisting antibodies we compared mumps specific serum IgG concentrations in two consecutive samples (*n* = 746), whereby the first sample was taken when students started their study prior to the outbreaks, and the second sample was taken 2-5 years later. We fit a binary mixture model to the data. The two mixing distributions represent uninfected and infected classes. Throughout we assume that the infection probability increases with the ratio of antibody concentrations of the second to first sample. The estimated infection attack rate is higher than reported earlier (0.095 versus 0.042). The analyses yield probabilistic classifications of participants, which are mostly quite precise owing to the high intraclass correlation in uninfected participants (0.85, 95%CrI: 0.82 − 0.87). The estimated probability of infection increases with decreasing antibody concentration in the pre-outbreak sample, such that the probability of infection is 0.12 (95%CrI: 0.10 − 0.13) for the lowest quartile of the pre-outbreak samples and 0.056 (95%CrI: 0.044 − 0.068) for the highest quartile. We discuss the implications of these insights for the design of booster vaccination strategies.

**Highlights:**

- We use paired pre- and post-outbreak serological data to estimate mumps infection rates in college students.
- We use a two-component mixture model to provide individual estimates of infection for each participant.
- The estimated population infection attack rate is higher than reported earlier (9.5% vs 4.2%).
- The estimated individual probability of infection increases with decreasing pre-outbreak antibody concentrations, from 12% in the lowest to 5.6% in the highest quartile.

## Introduction

Mass vaccination campaigns have been highly successful in reducing transmission and associated morbidity of infectious diseases of childhood. Case in point is the Measles-Mumps-Rubella vaccine, which in the Netherlands is administered at 14 months and 9 years and has a coverage of 90%*−*95%. It is known, however, that the antibody response to the mumps component in the vaccine wanes over time, and that IgG antibodies induced by the vaccine have relatively low avidity [1, 2, 3, 4]. Perhaps owing to this, mumps outbreaks have occurred worldwide over the past decades, mainly in vaccinated adolescents and young adults. These outbreaks often occurred in close contact settings (schools, households, parties), and were mostly caused by genotypes that are different from the vaccine genotype [5, 6, 7, 8, 9, 10, 11, 12, 13]. For instance, the current vaccine used in the Netherlands contains the Jeryl-Lynn strain (genotype A), and is genetically distant from the recent outbreak strains (genotypes D and G) [14, 15]. This vaccine is also widely used in the United States and other European countries.

Many but not all infections with mumps virus are asymptomatic or are associated with mild symptoms only, especially in vaccinated individuals [16]. This makes it difficult to assess the true extent of virus circulation in an outbreak. In principle, reliable infection attack rates can be obtained from measuring mumps-specific IgG antibody concentrations, because these generally increase after mumps virus infection [17]. However, a challenge is that paired pre- and post-outbreak samples are often not available, and that there are no generally agreed antibody concentrations that define recent infection in a single sample. Two studies have shown that pre-outbreak mumps neutralisation antibody concentrations in patients with mumps were generally lower than in persons who were not infected with mumps virus during the outbreak [2, 18, 19]. However, it proved not possible to define reliable cutoffs separating infected from uninfected persons, and separating patients with clinical symptoms from person with asymptomatic or mild infection [2]. Hence, it is not straight-forward to deduce who has been infected either clinically or sub-clinically, especially when only a single serum sample is available, and it is not clear how the probability of infection depends on pre-existing antibody concentrations.

In an earlier study, we measured mumps-specific IgG antibody concentrations in paired pre- and post-outbreak samples from university students in the Netherlands, using a fluorescent bead-based multiplex immunoassay [20, 21, 18]. In these studies, we calculated the proportions of symptomatic and asymptomatic infections and determined infection attack rates and risk factors for mumps virus infection using predefined criteria for infection. Specifically, participants would be classified as infected if there was a fourfold increase of antibody concentrations from first to second sample or high antibody concentration in the second sample. Further, to identify a correlate of protection, mumps-specific IgG concentrations in pre-outbreak samples were compared between infected and non-infected persons [22, 21, 23]. However, as antibody concentrations decay over time, especially in the first months after an infection [4, 24, 18, 19], and as the second sample in our study may have been taken several years after infection, we need analyses with less rigid criteria for infection. Therefore, we will in this study use methods that do not rely on predefined criteria for infection, but that derive informed probabilities of infection directly from the available serological data.

Specifically, to provide a probabilistic classification of participants we employ a two-component binary mixture model in which the component distributions represent infected and uninfected persons [25, 26, 27]. Usually, the mixing parameter in such analyses represents the prevalence or probability that a person is infected. Instead of using a fixed population-level mixing parameter, we here link the pre- and post-outbreak samples by making the biologically plausible assumption that the probability of infection increases monotonically with the ratio of post- to pre-outbreak antibody concentration. This enables estimation of the infection probability and associated uncertainty for each participant in a manner that is optimally informed by the data.

## Methods

### Study population and sample collection

All pre-outbreak sera in this retrospective study are taken from first-year medical students. Recruitment from this population was carried out from 2007-2010, and resulted in a study population of 746 students with both a pre- and post-outbreak sample. The study has been approved by a medical ethical committee (NL38042.041.11), and has been described in detail earlier [18]. Briefly, a self-sampled dried blot spot sample, a questionnaire concerning vaccination history, risk factors, and mumps symptoms has been obtained from each student. The serological criteria for mumps virus infection in the previous study were a fourfold increase in IgG concentration combined with at least an post-outbreak antibody concentration of 300 RU/ml [18]. A small subset of 16 participants show a strong (more than fourfold) decrease in the antibody concentration from pre- to post-outbreak sample. These could represent recent infections, as pre-outbreak samples had been taken from 2007-2010, while the outbreak lasted from 2009-2012. We therefore perform analyses of the full data in the body of the text, but also indicate how the results are affected if those 16 participants are left out.

### Mumps-specific IgG multiplex immunoassay

Mumps virus–specific serum immunoglobulin G (hereafter called IgG) antibody concentrations have been determined by a fluorescent bead–based multiplex immunoassay as described before, using purified Jeryl-Lynn mumps vaccine strain as antigen [20, 28]. For analysis, all antibody concentrations are log_2_-transformed, and in the following the log-transformed concentrations will be referred to as (antibody) titers.

### Mixture model for paired data

Participants are assumed to be uninfected in the first sample, and either infected or uninfected in the second sample. Here, we assume that these two classes are characterised by probability distributions for the antibody titers, and are characterised by density functions *f* ^uninf^ and *f* ^inf^. Based on preliminary analyses, we assume that the titer distributions of the two classes are normally distributed with parameters *θ*^uninf^ = {*μ*^uninf^, *σ*^uninf}^ and *θ*^inf^ = {*μ*^inf^, *σ*^inf}^. For our data, the pre-outbreak samples provide information on the uninfected component probability distribution, and the post-outbreak samples inform both the uninfected and infected component probability distributions. In mixture models for cross-sectional data the probability of infection is usually given by the prevalence weighted density of the infected component divided by the sum of the weighted infected and uninfected components (e.g., [27, 29, 30]). Here we take an alternative approach that makes use of the fact that data are paired, and in which we postulate that the probability that a post-outbreak sample belongs to the infected component (the mixing parameter) increases monotonically with the ratio of the post-versus pre-outbreak titers. Specifically, we propose a two-parameter logistic infection function such that the mixing parameter for the *i*-th participant (*i* = 1, …, 746), 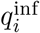, is given by

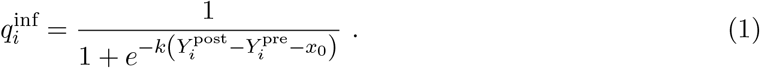

In equation (1), 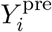 and 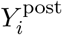 denote the antibody titers in the pre- and post-outbreak samples, *k* and *x*_0_ represent the steepness and 50% infection probability of the logistic function. Throughout, we use a transformation of the logistic function in terms of the probability that a sample belongs to the infected component at no increase in antibody concentration (*d*_0_) and fourfold increase in the antibody concentration (*d*_2_), as this yields parameters that can be interpreted more easily in biological terms. This, in turn, facilitates making informed choices for the parameter prior distributions. A straightforward calculation shows that

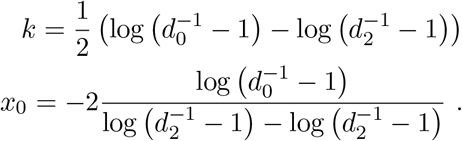

Biological reasoning implies that one would expect *d*_0_ *≈* 1 and *d*_2_ *≈* 1 [20, 21]. Notice further that 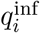 only depends on the difference of the two measurements but not on the individual values of 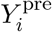 and 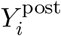. Also notice that a difference of 1 between antibody titers represents a twofold increase, a difference of 2 a fourfold increase, etcetera.

To estimate parameters and take correlations between pre- and post-outbreak samples into account, we estimate the unknown latent antibody titers of uninfected persons. Here we assume that the latent titers, 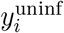, are drawn from a normal hyper distribution representing the unobserved true titers of uninfected persons. Specifically we take

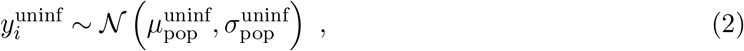

where 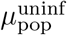 and 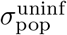 are the population mean and variance of the distribution of latent antibody concentrations in uninfected persons. Each measurement of uninfected persons provides a possibly imperfect representation of the latent antibody concentration, such that, for instance, for pre-outbreak samples 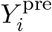 (*i* = 1, …, 746) we have

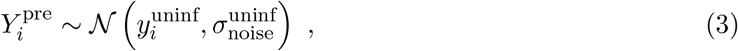

where 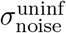 represents the measurement noise. On the other hand, post-outbreak samples can be either uninfected or infected, and therefore these samples are distributed according to the mixture distribution

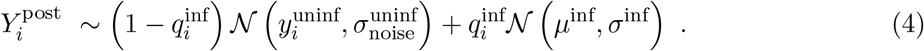

Together, equations (1) - (4) specify the model. Notice that the pre-outbreak samples inform the uninfected component distribution, and that the post-outbreak samples inform both the uninfected and infected component distributions.

With the above model at hand we can calculate for each participant the probability of infection, 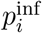 as the prevalence weighted density of the infected component divided by the sum of the weighted infected and uninfected components, i.e.

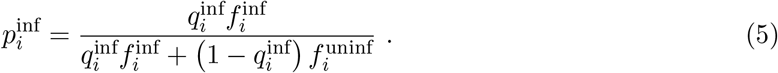

where we have suppressed the dependence of 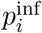 and 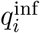 on the antibody differences 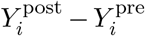 and the dependence of 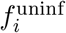 and 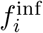 on post-outbreak antibody concentrations 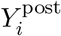. Hence, the probability of infection depends on both the mixing parameter and the mixing distributions.

Finally, to quantify correlations of antibody concentrations in pre- and post-outbreak samples of uninfected persons, we calculate the intraclass correlation *r*_ICC_ as

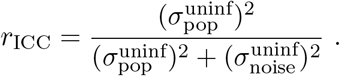

The intraclass correlation will be low if measurement noise is high and samples in uninfected persons are not strongly correlated, and it will be high if the reverse is true. Of course, whether post-outbreak samples are actually infected is not known, but will be estimated.

### Prior distributions and inference

Parameters are estimated in a Bayesian framework using Hamiltonian Monte Carlo, implemented in Stan [31]. The main parameters to be estimated are the mean and standard deviation of the hyper distribution of antibody titers in uninfected persons, 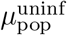 and 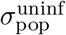, the random noise 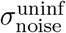, the mean and standard deviation of antibody titers in infected persons, *μ*^inf^ and *σ*^inf^, and the parameters defining the probability of infection when there is no increase in antibody titer from pre- to post-outbreak samples, and when there is a fourfold increase, *d*_0_ and *d*_2_.

Prior distributions for the means and standard deviations of the uninfected component distribution are based on pre-outbreak samples as these are by definition classified as uninfected. Specifically, we take 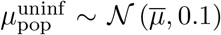 for the mean of the uninfected hyperdistribution and where 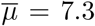 denotes the mean of the pre-outbreak antibody titers. Likewise, we take 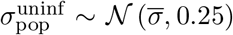 for the standard deviation of the uninfected hyperdistribution, where 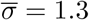 represents the standard deviation of the pre-outbreak data. Further, we take *d*_0_ *∼ ℬ* (1, 29) and *d*_2_ *∼ ℬ* (29, 1), such that the one-sided 95% prior ranges of these parameters are approximately [0, 0.1) and [0.9*−*1], respectively. Other parameters (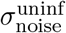, *μ*^inf^, and *σ*^inf^) are not equipped with explicit prior distributions, implying that all values on their domains are a priori equally likely.

All analyses are performed using R (version 4.3.1) and Stan using the RStan interface (version 2.21.8) [32]. We run 10 MCMC chains in parallel and base the analyses on 10,000 thinned samples from 10 well-mixed chains. Data, scripts, and figures are available in the online repository at https://github.com/rivm-syso/mumps_serology.

## Results

### Rule-based classification

The data have been described and analysed earlier [22, 21, 23, 18], and we here put the earlier findings into a perspective relevant to our analyses. Figure 1 shows the antibody titer measurements and distributions of the pre- and post-outbreak samples. There is substantial variation in the pre- and post-outbreak data, with mumps-specific IgG antibody titer measurements generally ranging from 4 *−* 14 (16 *−* 16, 384 RU/mL). Hence, antibody concentrations can vary more than 1, 000-fold. The figure also shows that there is a strong correlation between the paired pre- and post-outbreak samples, such that for the majority of participants the post-outbreak titer is roughly equal to the pre-outbreak titer. In fact, for most paired samples titer differences are within a twofold change in concentration. Only a small fraction of samples has a more than fourfold difference between pre- and post-outbreak samples, and most of these participants show a more than fourfold increase. In fact, 33*/*746 = 0.044 show a fourfold or more antibody increase. Of these, 31 also have a post-outbreak antibody concentration of 300 RU/mL, so that the rule-based cumulative infection attack rate would be 31*/*746 = 0.042. On the other hand, only a small fraction of participants (16*/*746 = 0.021) show a more than fourfold antibody decrease. Interestingly, the number of participants with a modest titer increase between two- and fourfold is substantially larger than the number with a similar modest decrease (56 versus 31). This suggests that some of the participants with modest titer increase may actually have been infected.

**Figure 1:**
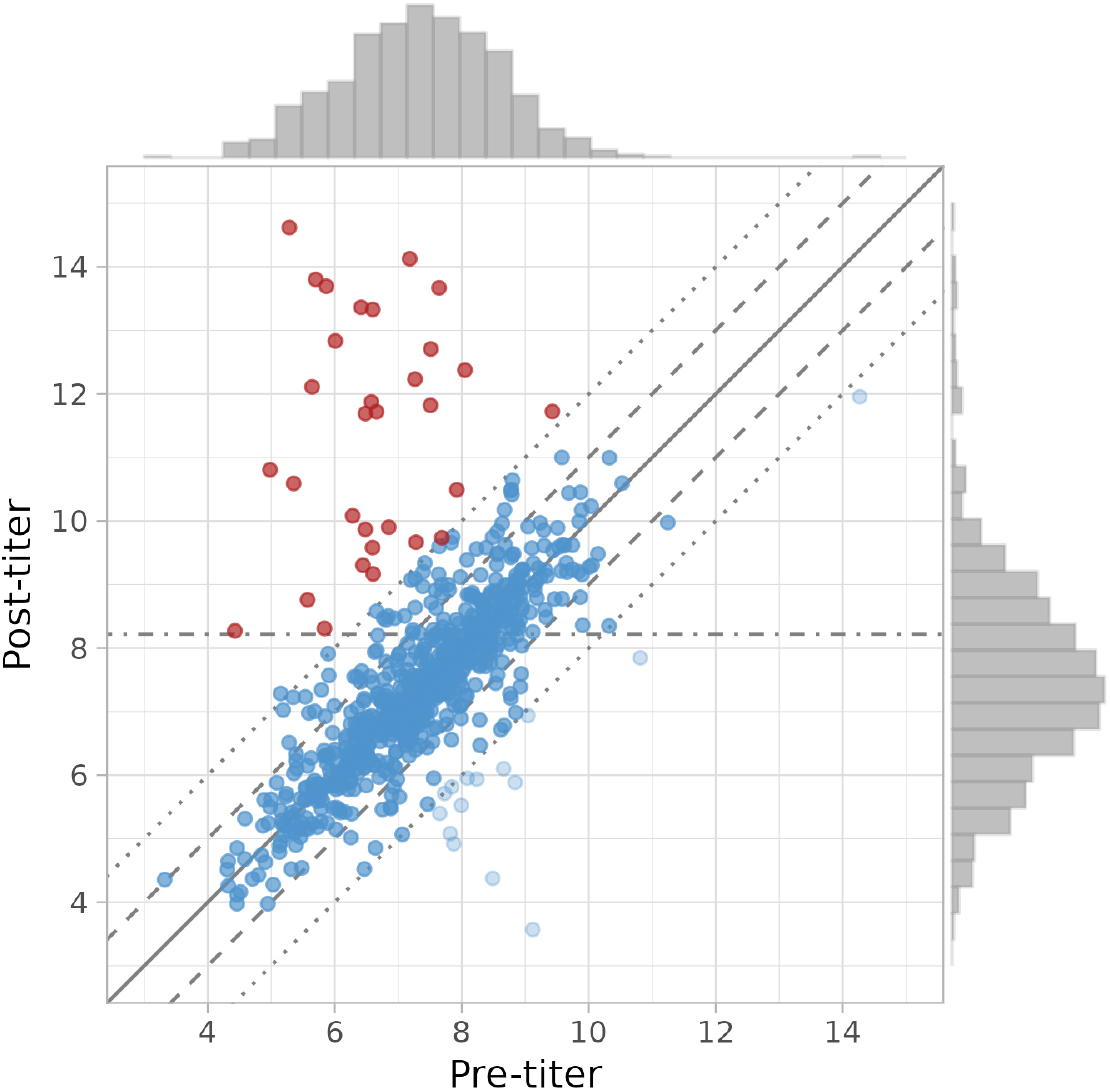
Overview of the data. Shown are the paired antibody titers (i.e. log_2_-transformed antibody concentrations) for each of the 746 participants in the post-versus pre-outbreak survey. Titers range from 3 to 15, so that mumps-specific IgG antibody concentrations range from 2^3^ = 8 RU/mL to 2^15^ = 32, 768 RU/mL [18]. The solid line represents the identity function with equal antibody concentrations in the pre- and post-outbreak samples. Dashed lines represent a twofold increase and decrease of the antibody concentration, and dotted lines represent a fourfold increase and decrease. The dashed-dotted line represents the minimum titer in the post-outbreak survey for positive classification (8.2, corresponding to 2^8.2^ = 300 RU/mL). Participants that were previously classified as infected are depicted in red [18]. Distributions on the top and right represent marginal titer distributions in the pre- and post-outbreak surveys, respectively.

In an earlier analyses a slightly different and more lenient classification rule for serological infection has been used [21]. Here, the authors opted for either a fourfold increase of antibody concentrations, or at least a post-outbreak titer of 10.6 (antibody concentration 1500 RU/mL). In this case, the number of infections and infection attack rates are 37 and 37*/*746 = 0.050, respectively.

### Parameter estimation

Next, we fit our mixture model (equations (1)-(4)) to the data. Parameter estimates and derived quantities are presented in Table 1, and the data and model fit are presented in Figure 2. Table 1 shows that the posterior mean of antibody concentrations in infected persons (9.89) is six times higher than the posterior mean in uninfected persons (7.28), hence 2^9.89*−*7.28^ = 6.1. However, the estimated antibody distributions are quite broad, especially for samples form uninfected persons. Overall, Figure 2 illustrates that there is a good correspondence between the model fit and data both for the pre- and post-outbreak surveys. Interestingly, even though there is considerable overlap between the mixing distributions of uninfected and infected persons, for most participants there is little doubt whether they had been infected or not. This is due to the high estimated intraclass correlation (0.85), and random variation in antibody concentrations of uninfected persons is estimated to be at most twofold 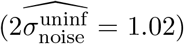. This, in turn, implies that already a twofold increase in antibody concentration provides evidence that the persons may have been infected.

**Table 1:** Parameter estimates and selected generated quantities. Estimates and generated quantities are represented by posterior medians and 95% posterior credible intervals. See text for explanation and details.

| Parameter | Description | Estimate | 95%CrI |
| --- | --- | --- | --- |
| $\mu_{\text{pop}}^{\text{uninf}}$ | Mean of antibody concentrations in uninfected participants | 7.28 | (7.20 – 7.36) |
| $\sigma_{\text{pop}}^{\text{uninf}}$ | Standard deviation of antibody concentrations in uninfected participants | 1.19 | (1.13 – 1.26) |
| $\sigma_{\text{noise}}^{\text{uninf}}$ | Random variation of antibody concentrations (e.g., measurement noise) | 0.51 | (0.48 – 0.54) |
| $\mu^{\text{inf}}$ | Mean of antibody concentrations in infected participants | 9.89 | (9.42 – 10.4) |
| $\sigma^{\text{inf}}$ | Standard deviation of antibody concentrations in infected participants | 1.91 | (1.63 – 2.27) |
| $d_0$ | Probability of infection at no antibody increase | 0.00034 | (0.000014 – 0.0025) |
| $d_1$ | Probability of infection at twofold antibody increase | 0.19 | (0.064 – 0.38) |
| $d_2$ | Probability of infection at fourfold antibody increase | 0.994 | (0.963 – 1.0) |
| $r_{\text{ICC}}$ | Intraclass correlation in uninfected participants | 0.85 | (0.82 – 0.87) |
| $p^{\text{inf}}$ | Overall probability of infection (cumulative incidence) | 0.095 | (0.082 – 0.11) |

**Figure 2:**
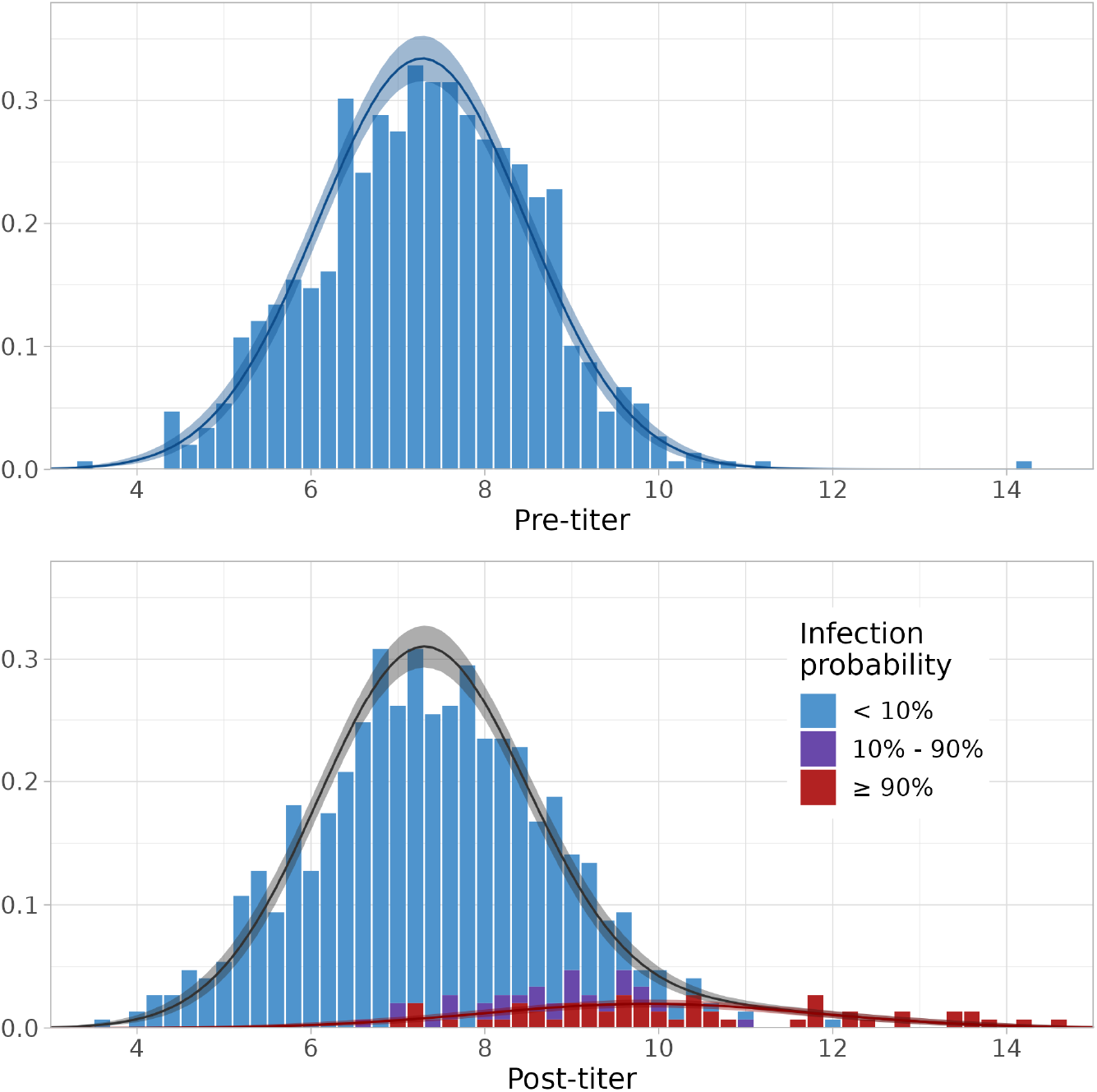
Overview of the data and fitted mixture distribution. Shown are the data (histograms) with fitted distributions weighted by estimated prevalence (lines). Shaded areas represent 95% credible envelopes. Top panel: pre-outbreak data with fitted distribution of uninfected participants. Bottom panel: post-outbreak data with fitted infected mixing distribution (red line) and overall mixture distribution (black line). Participants that are likely uninfected (posterior probability of infection ≤ 10%) are represented in blue, and those that are likely infected (posterior probability of infection ≥ 90%) are represented in red. Samples with intermediate posterior probabilities of infection are colored in purple.

In fact, the estimated probability of infection is essentially 0 if the post-outbreak titer equals the pre-outbreak titer (0.00034, 95%CrI: 0.000014 *−* 0.0025), is close to 1 in case of a fourfold antibody increase (0.994, 95%CrI: 0.963 *−* 1.0), and is almost 20% in case of a twofold increase (0.19, 95%CrI: 0.064 *−* 0.38). The overall estimated cumulative incidence of infection is estimated at 0.095 (95%CrI: 0.082 *−* 0.11) (Table 1).

To gauge the robustness of the above results with respect to the included participants, we rerun all analyses without the 16 participants with strongly decreased antibody concentrations. Overall, the results remain very similar, owing to the fact that these 16 only represent a small fraction of the total study population (*≈* 2%) while these participants are with high certainty classified as uninfected. Biggest difference is that the random variation from the latent antibody concentration in uninfected persons is estimated to be even smaller (0.37 versus 0.51), leading to a further increase in the intraclass correlation from 0.85 to 0.91. This in turn, leads to even higher precision with which individual probabilities of infection are estimated, and overall estimated cumulative incidence of infection is slightly increased (0.11, 95%CrI: 0.098 *−* 0.13).

Figure 3 shows 100 samples from the posterior distribution of the logistic infection function (equation (1)) together with estimated individual infections probabilities (equation (5)) as a function of the titer difference between pre- and post-outbreak samples. Most participants either have a post-outbreak titer that is close to the pre-outbreak titer, or have a post-outbreak titer that is substantially higher than the pre-outbreak titer. For these participants there is little doubt whether they have been infected or not. For instance, a titer difference of 0.5 translates to an increase of 41% of the antibody concentration (2^0.5^ = 1.4), and in this case it is highly unlikely that the increase has been caused by infection. Conversely, a difference of 1.6 translates to a threefold increase of the antibody concentration, and in this case it is approximately threefold more likely that the participant has been infected rather than that it has not been infected. Between these two extremes is the main uncertainty with respect to the infection status. Overall, there is a good correspondence between the infection function and individual estimated probabilities of infection, and this indicates that the model fits the data well.

**Figure 3:**
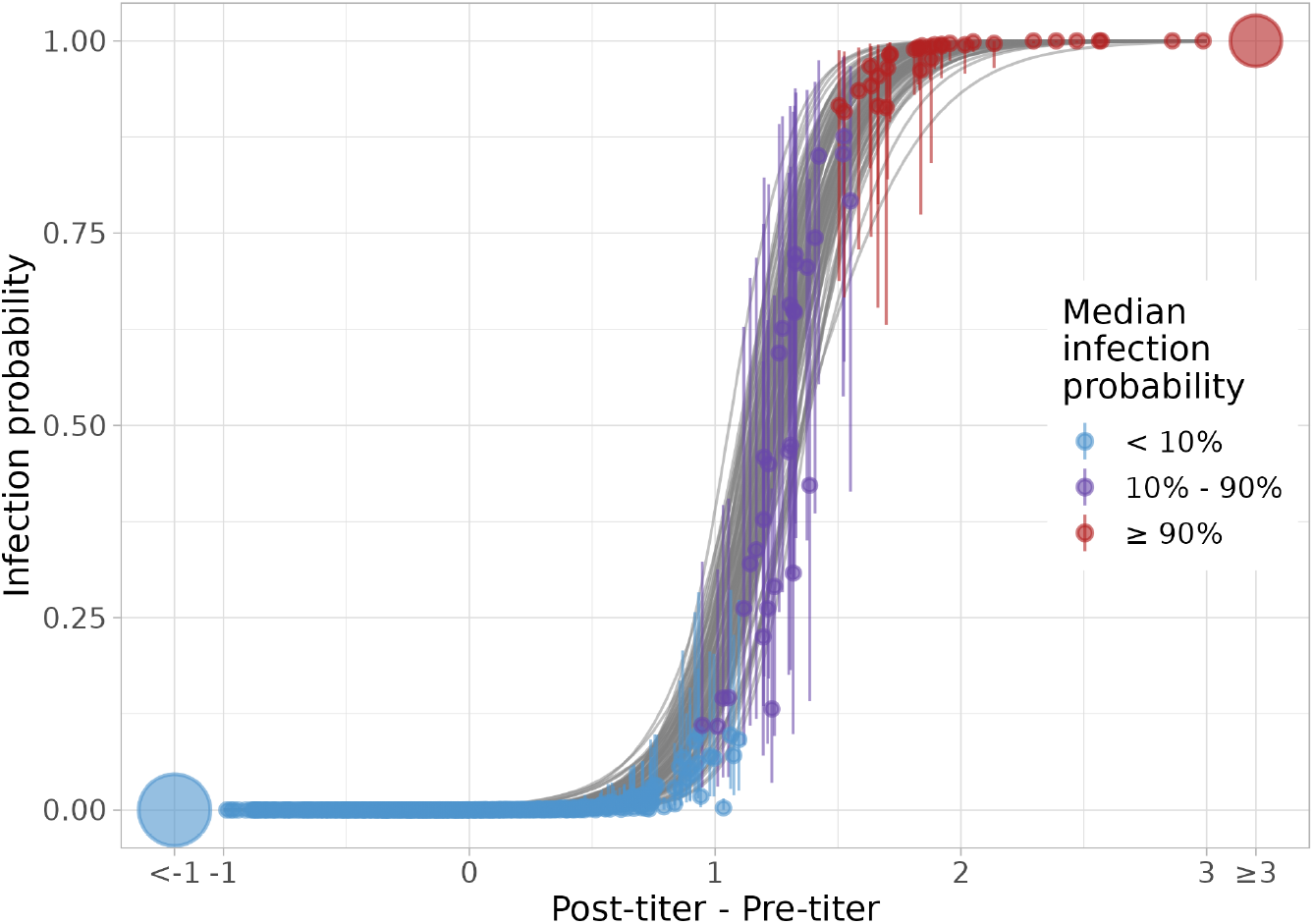
Estimated infection probabilities as function of differences in antibody concentrations. Shown are 100 samples from the posterior distribution of the logistic function determining the probability that a sample belongs to the infected component, as function of the difference in the antibody titers (grey lines). Also shown are the estimated (posterior median) infection probabilities for each of the 746 participants (dots), and uncertainty in the infection probabilities (95%CrI, whiskers). Notice that a difference of 1 between post- and pre-outbreak samples corresponds to a twofold increase, that a value of 2 corresponds to a fourfold increase, etcetera.

### Probability of infection as function of pre-outbreak antibody titer

Next, we turn attention to the relation between pre-outbreak antibody titers and subsequent probability of infection. Figure 4 shows for all 746 participants the estimated infection probability with associated uncertainty as function of the pre-outbreak titer. In general, there is limited uncertainty for most participants, especially when the estimated probability of infection is close to 0 or close to 1. Only for a small subset of 31 participants with estimated posterior median probability of infection ≥ 10% and *<* 90% is there substantial uncertainty.

**Figure 4:**
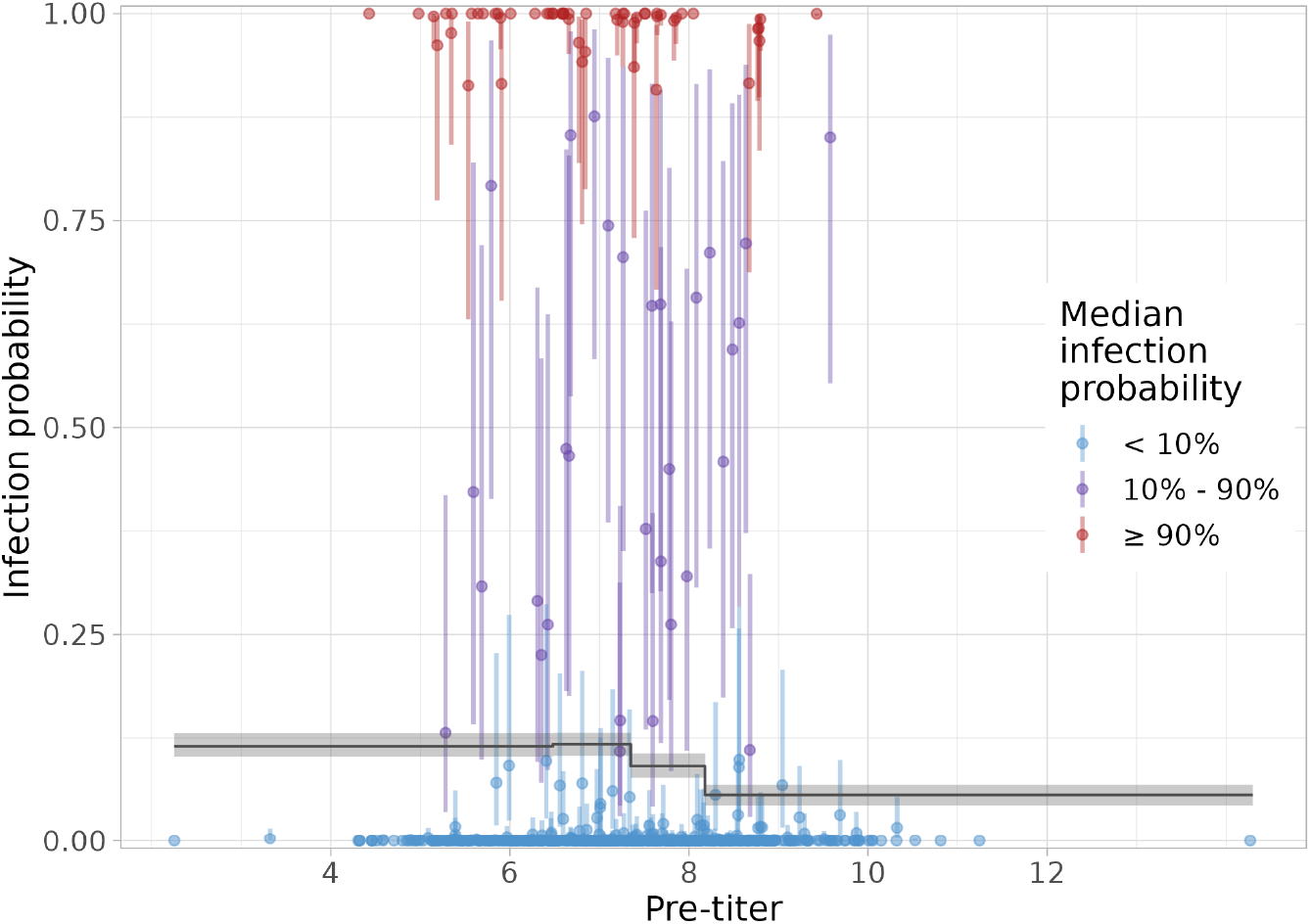
Estimated infection probabilities as function of the pre-outbreak antibody titer. Shown are the estimated infection probabilities for all participants (dots: posterior medians; lines: 95% credible interval). Colors represent three infection probability classes (cf. Figure 2). Also shown are estimated infection probabilities (median and 95% credible intervals), stratified by quartile of pre-outbreak antibody concentration.

To generalise these results for groups of persons and estimate the probability of infection as function of the pre-outbreak antibody titer we have stratified the study population by pre-outbreak antibody titers and calculated the probabilities of infection in these groups. Specifically, we have grouped pre-outbreak titers in four equal sized groups, with titer ranges [2, 6.48), [6.48, 7.35), [7.35, 8.18), and [8.18, 15), corresponding to antibody concentrations of 4 *−* 89 RU/mL, 89 *−* 163 RU/mL, 163 *−* 290 RU/mL, and 290 *−* 32, 768 RU/mL. As it appears, the estimated probabilities of infection generally decrease with increasing pre-outbreak antibody titer. Specifically, the estimated infection probability is 0.12 (95%CrI: 0.10 *−* 0.13) for the lowest two quartiles, and 0.056 (95%CrI: 0.044 *−* 0.068) for the highest quartile. Hence, while there does not appear to be a fixed antibody titer that provides protection against infection, the probability of infection is more than double in the groups with low pre-outbreak antibody titers as compared to the group with highest antibody titers. The estimated overall probability of infection is 0.95 (95%CrI: 0.082 *−* 0.11) (1). Thus, our results confirm earlier results that the probability of infection increases with decreasing antibody titers [18], but yield substantially higher infection attack rates than reported earlier.

## Discussion

The past two decades have witnessed a distinct increase in the number of outbreaks of mumps in highly vaccinated populations, mostly by non-vaccine genotypes, with an over-representation of adolescents and young adults, and predominantly in close-contact settings (households, schools and universities, parties) [33]. This has prompted suggestions that a booster vaccination in adolescence could be beneficial ([18, 33] and references therein). Indeed, limited experience with such booster doses has largely been positive [34, 33, 24, 18]. Our analyses have provided additional support for the potential benefit of an extra vaccination in this age group to improve immunity to mumps. Here, we have shown that the infection attack rate can be high in Dutch university university (9.5%), and that in addition the infection attack rate is more than twice as high in students with low pre-existing mumps-specific IgG antibody concentrations than in students with high pre-existing antibodies (12% vs 5.6%). This suggests that the hazard of infection can be at least twice as high in adolescents and young adults with low antibody concentrations than in those with high antibody concentrations. However, no fixed pre-outbreak antibody concentration could be reliably determined that fully correlates with protection against infection. This indicates that not only IgG antibodies but also other compartments of the immune system, such as cellular immunity, play a role in protection against mumps [35, 36]. Nevertheless, young adults with low mumps-specific IgG antibody concentrations are at higher risk for infection and may therefore benefit most from a third dose of the MMR vaccine. Interestingly in this context, our previous study showed that individuals with lowest pre-vaccination IgG concentrations also showed the strongest increase in IgG concentrations after vaccination [24].

Our analyses build on and extend earlier analyses for the same outbreak [21, 18]. An advantage of the current analyses is that they are fully self-contained, and that all parameters, distributions, and infection rates are directly estimated from the data. This is a considerable advantage and makes the analyses quite robust, as there is no fixed predefined level of antibody concentrations that is indicative of infection or protection against infection [37, 38]. Moreover, antibody concentrations in the population depend in a complex manner on time since last vaccination, potential previous infection(s), and outbreak genotype(s). Therefore, it is unlikely that proper sets of validation samples with confirmed uninfected and infected persons can be obtained. This is true not only for our population of university students but probably holds more generally. Thus, ad hoc choices have previously been made for the infection criterion (a fourfold antibody increase with additional requirements depending on the study). Comparing our results with the earlier analyses of Gouma et al. [21] and Kaaijk et al. [18] we find a similar pattern that infection attack rates increase with decreasing prior antibody concentrations. Our analyses also indicate, however, that infection attack rates may have been substantially higher (9.5%) than reported earlier (4% *−* 6%). This is due to the fact that many participants with modest increases of antibody titers (two- to threefold) may actually have been infected. In all, our results paint a more dynamic picture of the antibody dynamics induced by infection and waning than hitherto considered.

Previous analyses for bivariate serological cross-sectional studies [26, 39, 40] have modelled the serological response with bivariate mixture distributions. Here, by virtue of the fact that we have paired data at our disposal we have been able to couple pre- and post-outbreak data using an infection function (equation (1)). This has enabled precise estimation of infection probabilities for the majority of participants even though the distributions of uninfected and infected persons show considerable overlap. In fact, we would not have been able to obtain precise estimates of infection probabilities if the data had been treated as two separate cross-sectional surveys (J. Gomme, unpublished report). In principle, our model and estimates can also be used for prediction of infection probabilities of paired samples in future studies that have been analysed with the test employed here.

In our analyses we have included 16 participants with a strong (at least fourfold) antibody decrease. These person may have had experienced a recent mumps infection, and may therefore not be representative for antibody concentrations in populations with no recent exposure. To study how the results are affected by this choice, we have performed a sensitivity analysis in which these samples are excluded. Fortunately, the results are largely unaffected by inclusion of exclusion of those participants. This due to the fact that these 16 persons represent only a small minority of the study population (2%), and the fact that the estimated probability of infection for those participants is essentially 0.

Two main limitations deserve scrutiny. First, we have throughout focused on probabilistic classification of participants and estimation of the serological infection attack rate. It would be desirable, but not possible with the limited information available in our study, if the analyses could be extended to include not only serological data but also information on the severity of infection. It would be particularly interesting to study how the severity of infection depends on both the pre- and post-outbreak antibody concentrations. Second, our analyses apply to the specific situation of medical students in the Netherlands. This is a very specific population and setting. For instance, in the Netherlands the vast majority of students have been vaccinated, and contact intensities in this group are probably substantially higher than in other strata of the population of similar age. It remains therefore an open question to which extent the results still apply in other settings. In particular, one could envisage that our estimated infection attack rates in medical students represent an upper bound for this age group in general.

Finally, analyses for paired data such as presented here may have wider applicability, and are not restricted to serological surveys for infectious diseases. Indeed, our methods of analysis apply whenever a latent response is usually stable but highly individual-specific. In such instances, it may be of interest to be able to decide when there are substantial deviations from this stable individual-specific response. Such situations occur often in hospital settings when patients are regularly screened for the onset of disease. Our study has shown one way how such data can be analysed elegantly with mixture model-based methods.

## Data Availability

All data produced are available online at https://github.com/rivm-syso/mumps_serology

## Acknowledgements

We gratefully acknowledge the participants of the study. Part of this study has been performed during an internship of JG from the Department of Epidemiology and Social Medicine at University of Antwerp at the National Institute of Public Health and the Environment (RIVM). This study was supported by the Dutch Ministry of Health, Welfare and Sport.

## Notes

### Competing Interest Statement

The authors have declared no competing interest.

### Funding Statement

This study was funded by the Dutch Ministry of Health, Welfare and Sport.

### Author Declarations

Data used in this study have been published and described previously (Kaaijk et al 2020, J Infect Dis 221, 902-909), and approval had previously been given by a medical ethical committee (NL38042.041.11).

## References

[1] I. Davidkin, S. Jokinen, M. Broman, P. Leinikki, and H. Peltola. Persistence of measles, mumps, and rubella antibodies in an MMR-vaccinated cohort: a 20-year follow-up. J Infect Dis, 197(7):950–956, Apr 2008.

[2] M. M. Cortese, A. E. Barskey, G. E. Tegtmeier, C. Zhang, L. Ngo, M. H. Kyaw, A. L. Baughman, J. E. Menitove, C. J. Hickman, W. J. Bellini, G. H. Dayan, G. R. Hansen, and S. Rubin. Mumps antibody levels among students before a mumps outbreak: in search of a correlate of immunity. J Infect Dis, 204(9):1413–1422, Nov 2011.

[3] M. Kontio, S. Jokinen, M. Paunio, H. Peltola, and I. Davidkin. Waning antibody levels and avidity: implications for MMR vaccine-induced protection. J Infect Dis, 206(10):1542–1548, Nov 2012.

[4] A. Antia, H. Ahmed, A. Handel, N. E. Carlson, I. J. Amanna, R. Antia, and M. Slifka. Heterogeneity and longevity of antibody memory to viruses and vaccines. PLoS Biol, 16(8):e2006601, Aug 2018.

[5] M. van Boven, W. L. Ruijs, J. Wallinga, P. D. O’Neill, and S. é. Estimation of vaccine efficacy and critical vaccination coverage in partially observed outbreaks. PLoS Comput Biol, 9(5):e1003061. 2013.

[6] L. J. Willocks, D. Guerendiain, H. I. Austin, K. E. Morrison, R. L. Cameron, K. E. Templeton, V. R. F. DE Lima, R. Ewing, W. Donovan, and K. G. J. Pollock. An outbreak of mumps with genetic strain variation in a highly vaccinated student population in Scotland. Epidemiol Infect, 145(15):3219–3225, Nov 2017.

[7] L. N. Patel, R. J. Arciuolo, J. Fu, F. R. Giancotti, J. R. Zucker, J. L. Rakeman, and J. B. Rosen. Mumps Outbreak Among a Highly Vaccinated University Community-New York City, January-April 2014. Clin Infect Dis, 64(4):408–412, Feb 2017.

[8] L. Veneti, K. Borgen, K. S. Borge, K. Danis, M. Greve-Isdahl, K. Konsmo, G. lstad S. A. ø, K. S. ystese, R. Rykkvin, E. Sagvik, and R. Riise. Large outbreak of mumps virus genotype G among vaccinated students in Norway, 2015 to 2016. Euro Surveill, 23(38), Sep 2018.

[9] M. Shah, P. Quinlisk, A. Weigel, J. Riley, L. James, J. Patterson, C. Hickman, P. A. Rota, R. Stewart, N. Clemmons, N. Kalas, C. Cardemil, M. Patel, M. Donahue, A. Schneider, U. Ukegbu, K. Wittich, J. Kellogg, D. Beardsley, N. Tran, D. Callaghan, A. Pyatt, T. Kitzmann, and B. Kintigh. Mumps Outbreak in a Highly Vaccinated University-Affiliated Setting Before and After a Measles-Mumps-Rubella Vaccination Campaign-Iowa, July 2015-May 2016. Clin Infect Dis, 66(1):81–88, Jan 2018.

[10] D. W. Westphal, A. Eastwood, A. Levy, J. Davies, C. Huppatz, M. Gilles, H. Lyttle, S. A. Williams, and G. K. Dowse. A protracted mumps outbreak in Western Australia despite high vaccine coverage: a population-based surveillance study. Lancet Infect Dis, 19(2):177–184, Feb 2019.

[11] A. Ferenczi, S. Gee, S. Cotter, and K. Kelleher. Ongoing mumps outbreak among adolescents and young adults, Ireland, August 2018 to January 2020. Euro Surveill, 25(4), Jan 2020.

[12] M. Perez Duque, A. San-Bento L. on, P. dio M. A. a, M. J. Albuquerque, M. Nascimento, S. Balasegaram, and R. Machado. Mumps outbreak among fully vaccinated school-age children and young adults, Portugal 2019/2020. Epidemiol Infect, 149:e205, Aug 2021.

[13] L. H. Moncla, A. Black, C. DeBolt, M. Lang, N. R. Graff, A. C. rez Osorio, N. F. ller, D. Haselow, S. Lindquist, and T. Bedford. Repeated introductions and intensive community transmission fueled a mumps virus outbreak in Washington State. Elife, 10, Apr 2021.

[14] S. A. Rubin, M. A. Link, C. J. Sauder, C. Zhang, L. Ngo, B. K. Rima, and W. P. Duprex. Recent mumps outbreaks in vaccinated populations: no evidence of immune escape. J Virol, 86(1):615–620, Jan 2012.

[15] S. Gouma, T. Vermeire, S. Van Gucht, L. Martens, V. Hutse, J. Cremer, P. A. Rota, G. Leroux-Roels, M. Koopmans, R. van Binnendijk, and E. Vandermarliere. Differences in antigenic sites and other functional regions between genotype A and G mumps virus surface proteins. Sci Rep, 8(1):13337, Sep 2018.

[16] S. Dittrich S. é, A. van Lier, R. Kohl, H. Boot, M. Koopmans, and R. van Binnendijk. Assessment of serological evidence for mumps virus infection in vaccinated children. Vaccine, 29(49):9271–9275, Nov 2011.

[17] S. Borgmann, F. Schwab, S. Santibanez, and A. Mankertz. Mumps virus infection in vaccinated patients can be detected by an increase in specific IgG antibodies to high titres: a retrospective study. Epidemiol Infect, 142(11):2388–2396, Nov 2014.

[18] Patricia Kaaijk, Alienke J Wijmenga-Monsuur, Marlies A van Houten, Irene K Veldhuijzen, Hinke I ten Hulscher, Jeroen Kerkhof, Fiona R van der Klis, and Rob S van Binnendijk. A Third Dose of Measles-Mumps-Rubella Vaccine to Improve Immunity Against Mumps in Young Adults. The Journal of Infectious Diseases, 221(6):902– 909, 04 2019.

[19] Patricia Kaaijk, Alienke J. Wijmenga-Monsuur, Hinke I. ten Hulscher, Jeroen Kerkhof, Gaby Smits, Mioara Alina Nicolaie, Marianne A. van Houten, and Rob S. van Binnendijk. Antibody levels at 3-years follow-up of a third dose of measles-mumps-rubella vaccine in young adults. Vaccines, 10(1), 2022.

[20] G. P. Smits, P. G. van Gageldonk, L. M. Schouls, F. R. van der Klis, and G. A. Berbers. Development of a bead-based multiplex immunoassay for simultaneous quantitative detection of IgG serum antibodies against measles, mumps, rubella, and varicella-zoster virus. Clin Vaccine Immunol, 19(3):396–400, Mar 2012.

[21] S. Gouma, T. M. Schurink-Van’t Klooster, H. E. de Melker, J. Kerkhof, G. P. Smits S. J. é, C. A. van Els, G. J. Boland, A. C. Vossen, P. R. Goswami, M. P. Koopmans, and R. S. van Binnendijk. Mumps serum antibody levels before and after an outbreak to assess infection and immunity in vaccinated students. Open Forum Infect Dis, 1(3):ofu101, Dec 2014.

[22] J. Sane, S. Gouma, M. Koopmans, H. de Melker, C. Swaan, R. van Binnendijk, and S. é. Epidemic of mumps among vaccinated persons, The Netherlands, 2009-2012. Emerg Infect Dis, 20(4):643–648, Apr 2014.

[23] S. Gouma, J. Sane, D. Gijselaar, J. Cremer S. é, M. Koopmans, and R. van Binnendijk. Two major mumps genotype G variants dominated recent mumps outbreaks in the Netherlands (2009-2012). J Gen Virol, 95(Pt 5):1074–1082, May 2014.

[24] Patricia Kaaijk, M Alina Nicolaie, Debbie van Rooijen, Marianne A van Houten, Fiona R van der Klis, Anne-Marie Buisman, and Rob S van Binnendijk. Dynamics of the Antibody Response After a Third Dose of Measles-Mumps-Rubella Vaccine Indicate a Slower Decline Compared With a Second Dose. Open Forum Infectious Diseases, 7(11):ofaa505. 10 2020.

[25] A. Steens, S. Waaijenborg, P. F. Teunis, J. H. Reimerink, A. Meijer, M. van der Lubben, M. Koopmans, M. A. van der Sande, J. Wallinga, and M. van Boven. Age-dependent patterns of infection and severity explaining the low impact of 2009 influenza A (H1N1): evidence from serial serologic surveys in the Netherlands. Am J Epidemiol, 174(11):1307–1315, Dec 2011.

[26] D. te Beest, E. de Bruin, S. Imholz, J. Wallinga, P. Teunis, M. Koopmans, and M. van Boven. Discrimination of influenza infection (A/2009 H1N1) from prior exposure by antibody protein microarray analysis. PLoS One, 9(11):e113021, 2014.

[27] M. A. Vink, J. van de Kassteele, J. Wallinga, P. F. Teunis, and J. A. Bogaards. Estimating seroprevalence of human papillomavirus type 16 using a mixture model with smoothed age-dependent mixing proportions. Epidemiology, 26(1):8–16, Jan 2015.

[28] G. Smits, L. Mollema, H. de Melker, I. Tcherniaeva, S. Waaijenborg, R. van Binnendijk, F. van der Klis, and G. Berbers. Seroprevalence of mumps in The Netherlands: dynamics over a decade with high vaccination coverage and recent outbreaks. PLoS One, 8(3):e58234, 2013.

[29] J. A. Bouman, J. Riou, S. Bonhoeffer, and R. R. Regoes. Estimating the cumulative incidence of SARS-CoV-2 with imperfect serological tests: Exploiting cutoff-free approaches. PLoS Comput Biol, 17(2):e1008728, Feb 2021.

[30] J. A. Bouman, S. Kadelka, S. Stringhini, F. Pennacchio, B. Meyer, S. Yerly, L. Kaiser, I. Guessous, A. S. Azman, S. Bonhoeffer, and R. R. Regoes. Applying mixture model methods to SARS-CoV-2 serosurvey data from Geneva. Epidemics, 39:100572, Jun 2022.

[31] B. Carpenter, A. Gelman, M. Hoffman, D. Lee, B. Goodrich, M. Betancourt, M. Brubaker, J. Guo, P. Li, and A. Riddell. Stan: A probabilistic programming language. Journal of Statistical Software, Articles, 76(1):1–32, 2017.

[32] Bob Carpenter, Andrew Gelman, Matthew D. Hoffman, Daniel Lee, Ben Goodrich, Michael Betancourt, Marcus Brubaker, Jiqiang Guo, Peter Li, and Allen Riddell. Stan: A probabilistic programming language. Journal of Statistical Software, 76(1):1–32, 2017.

[33] E. Lam, J. B. Rosen, and J. R. Zucker. Mumps: an Update on Outbreaks, Vaccine Efficacy, and Genomic Diversity. Clin Microbiol Rev, 33(2), Mar 2020.

[34] C. V. Cardemil, R. M. Dahl, L. James, K. Wannemuehler, H. E. Gary, M. Shah, M. Marin, J. Riley, D. R. Feikin, M. Patel, and P. Quinlisk. Effectiveness of a Third Dose of MMR Vaccine for Mumps Outbreak Control. N Engl J Med, 377(10):947–956, Sep 2017.

[35] S. Jokinen, P. Osterlund, I. Julkunen, and I. Davidkin. Cellular immunity to mumps virus in young adults 21 years after measles-mumps-rubella vaccination. J Infect Dis, 196(6):861–867, Sep 2007.

[36] J. de Wit, M. E. Emmelot, M. C. M. Poelen, R. S. van Binnendijk, S. van der Lee, D. van Baarle, W. G. H. Han, C. A. C. M. van Els, and P. Kaaijk. T-cell memory. J Allergy Clin Immunol, 141(5):1908–1911, May 2018.

[37] P. F. Teunis, J. C. van Eijkeren, W. F. de Graaf A. B. ć, and M. E. Kretzschmar. Linking the seroresponse to infection to within-host heterogeneity in antibody production. Epidemics, 16:33–39, Sep 2016.

[38] P. F. M. Teunis and J. C. H. van Eijkeren. Estimation of seroconversion rates for infectious diseases: Effects of age and noise. Stat Med, 39(21):2799–2814, Sep 2020.

[39] D. E. Te Beest, P. J. Birrell, J. Wallinga, D. De Angelis, and M. van Boven. Joint modelling of serological and hospitalization data reveals that high levels of pre-existing immunity and school holidays shaped the influenza A pandemic of 2009 in the Netherlands. J R Soc Interface, 12(103), Feb 2015.

[40] M. A. Vink, J. Berkhof, J. van de Kassteele, M. van Boven, and J. A. Bogaards. A Bivariate Mixture Model for Natural Antibody Levels to Human Papillomavirus Types 16 and 18: Baseline Estimates for Monitoring the Herd Effects of Immunization. PLoS One, 11(8):e0161109, 2016.

